# Segmental Signs and Spontaneous Pain in Acute Visceral Disease - Lateralization and Bodily Patterns

**DOI:** 10.1101/2020.07.23.20160598

**Authors:** Nour Shaballout, Anas Aloumar, Jorge Manuel, Marcus May, Florian Beissner

## Abstract

The differential diagnosis of acute visceral diseases is a challenging clinical problem. The older literature suggests that patients with acute visceral problems show segmental signs, such as hyperalgesia, skin resistance, or muscular defence, whose lateralization and segmental distribution may be used for differential diagnosis.

This study aimed to investigate the lateralization and segmental distribution of spontaneous pain and segmental signs in acute visceral diseases.

We recruited 208 emergency room patients that were presenting for acute medical problems. All patients underwent a structured 10-minute bodily examination to test for various segmental signs and were asked for spontaneous pain and segmental symptoms, such as nausea, meteorism, and urinary retention. We collected all findings as digital drawings on a tablet-PC. After the final diagnosis, patients were divided into groups according to the organ affected. Using statistical image analysis, we calculated average distributions of pain and segmental signs for the heart, lungs, stomach, liver/gallbladder, and kidneys/ureters analyzing their segmental distribution and lateralization.

85 of 110 patients with a single-organ problem reported pain, while 81 had at least one segmental sign, the most frequent being hyperalgesia (n=46), and muscle resistance (n=39). While the pain was distributed along the body midline, segmental signs for the heart, stomach and liver/gallbladder appeared mostly ipsilateral to the affected organ. An unexpectedly high number of patients (n=37) further showed ipsilateral mydriasis.

The present study underlines the usefulness of including segmental signs in the bodily examination of patients with acute medical problems.

## Introduction

The differential diagnosis of acute visceral diseases is a common but challenging clinical problem. Since pain originating from visceral organs often exhibits characteristic patterns [1-8], many textbooks assign pain location a discriminative role in the differential diagnosis. However, many studies have also reported negative results when testing the predictive power of pain location [9-10]. For example, pain localization in patients with coronary heart disease (CHD) does not significantly differ from non-CHD chest pain patients [11].

Closely related but much less known phenomena in acute visceral diseases are segmental signs and symptoms [12-27]. Here, pain signals from visceral organs are referred to other somatic or visceral tissues with overlapping segmental innervation. Such pain signals most frequently manifest themselves in the form of referred hyperalgesia of the skin, a phenomenon first described by Ross and Sturge in the 1880s [28-29] and subsequently studied in depth by Head and Mackenzie [12-15]. Head mapped out the cutaneous zones of referred hyperalgesia for all major organs and compared them to the location of skin lesions in herpes zoster [12]. The result is still considered one of the most precise maps of segmental innervation [30-31].

To the present day zones of referred hyperalgesia in visceral disease carry Head’s name in many European countries, such as France, Germany, and Spain. In other parts of the world, however, the term “Head’s zones”, as well as Head’s work in general, is hardly known. Some authors even speak of segmental anatomy as a “wrongly forgotten science” [26]. Still less known is the fact that referred signs are not limited to hyperalgesia of the skin but instead show a plethora of manifestations, including sensory disturbances, like allodynia, and deep hyperalgesia (also known as Mackenzie’s zones), motor disturbances, like increased resistance of the skin, muscular defence, and resistance to passive joint movement, and, finally, signs of sympathetic activation, like vasomotor changes, anisohydrosis, piloerection, and even anisocoria. Furthermore, segmental signs may be accompanied by symptoms of viscero-visceral reflexes, such as nausea, vomiting, diarrhoea, constipation, meteorism, and urinary retention.

To our knowledge, a systematic evaluation of simultaneously collected segmental signs and symptoms in patients has never been published in the English scientific literature. In Germany, however, Karl Hansen (1893–1962) and Hans Schliack (1919–2008) have studied a wide variety of segmental signs over several decades. While their results have only been published in German [31], the essence of their work has recently been made available in book form and extended by the work of other clinicians [26]. In a large sample of internal medicine patients, Hansen and Schliack confirmed many of Head’s observations and greatly extended them to include all of the above-mentioned segmental signs and symptoms [31]. Even more than Head, the authors emphasized the importance of sign lateralization by defining a side rule according to which segmental signs are most likely to appear ipsilateral to the affected organ (**Table 1**).

**Table 1:** Lateralization of segmental signs for individual organs according to Hansen & Schliack [31].

| right | left |
| --- | --- |
| (Heart) | Heart |
| - | Pericardium |
| (Aorta) | Aorta |
| Right lung and bronchi | Left lung and bronchi |
| Right pleura | Left pleura |
| - | Stomach (corpus, fundus) |
| Pylorus | - |
| Duodenum | - |
| - | Jejunum |
| Ileum | - |
| - | Pancreas |
| Liver | - |
| Gallbladder | - |
| - | Spleen |
| Caecum | - |
| Appendix | - |
| Colon asc. | - |
| Colon transv. prox. part | - |
| - | Colon transv. dist. part |
| - | Colon desc. |
| - | Sigmoid colon |
| - | Rectum |
| Right kidney | Left kidney |
| Right ureter | Left ureter |
| Right testis, ovary, salpinx | Left testis, ovary, salpinx |

A methodological problem that has hampered clinical research on segmental signs for a long time is the difficulty in adequately measuring bodily signs. However, recent developments in the field of digital pain drawings offer new and exciting possibilities to systematically record segmental signs and analyze them using methods of statistical image analysis [32].

Here, we report the results of a study that investigated both the bodily patterns and lateralizations of segmental signs and spontaneous pain in acute visceral diseases. We aimed to derive average distributions of spontaneous pain and segmental signs for as many internal organs as possible and to analyze their segmental content and lateralization. To achieve this, we combined digital pain drawing technology and a structured 10-minutes bodily examination in patients presenting to the emergency room.

## Materials and methods

### Ethics

The study was approved by the Ethics Committee of Hannover Medical School (#2987-2017) and conducted under the Declaration of Helsinki. All patients gave written informed consent after being informed about the purpose of the study.

### Study population

Our study population consisted of patients from the emergency department of Hannover Medical School, who were referred to the internal medicine physicians. Eligible patients were adults (age ≥ 18 years in Germany) presenting with an acute medical problem and with the ability to give written informed consent. Furthermore, patients needed to be oriented as to place, time, and person. Exclusion criteria comprised refusal or inability to provide written consent, previously known or acutely diagnosed spinal cord injury, pregnancy, acute or past illnesses which (in the investigator’s opinion) could negatively affect the outcome of the study (e.g., known ocular or peripheral nervous disease), uncooperative patients, and patients who only presented to the emergency room for educational purposes or to receive a prescription. For a flow-chart, see **Figure 1**.

**Figure 1:**
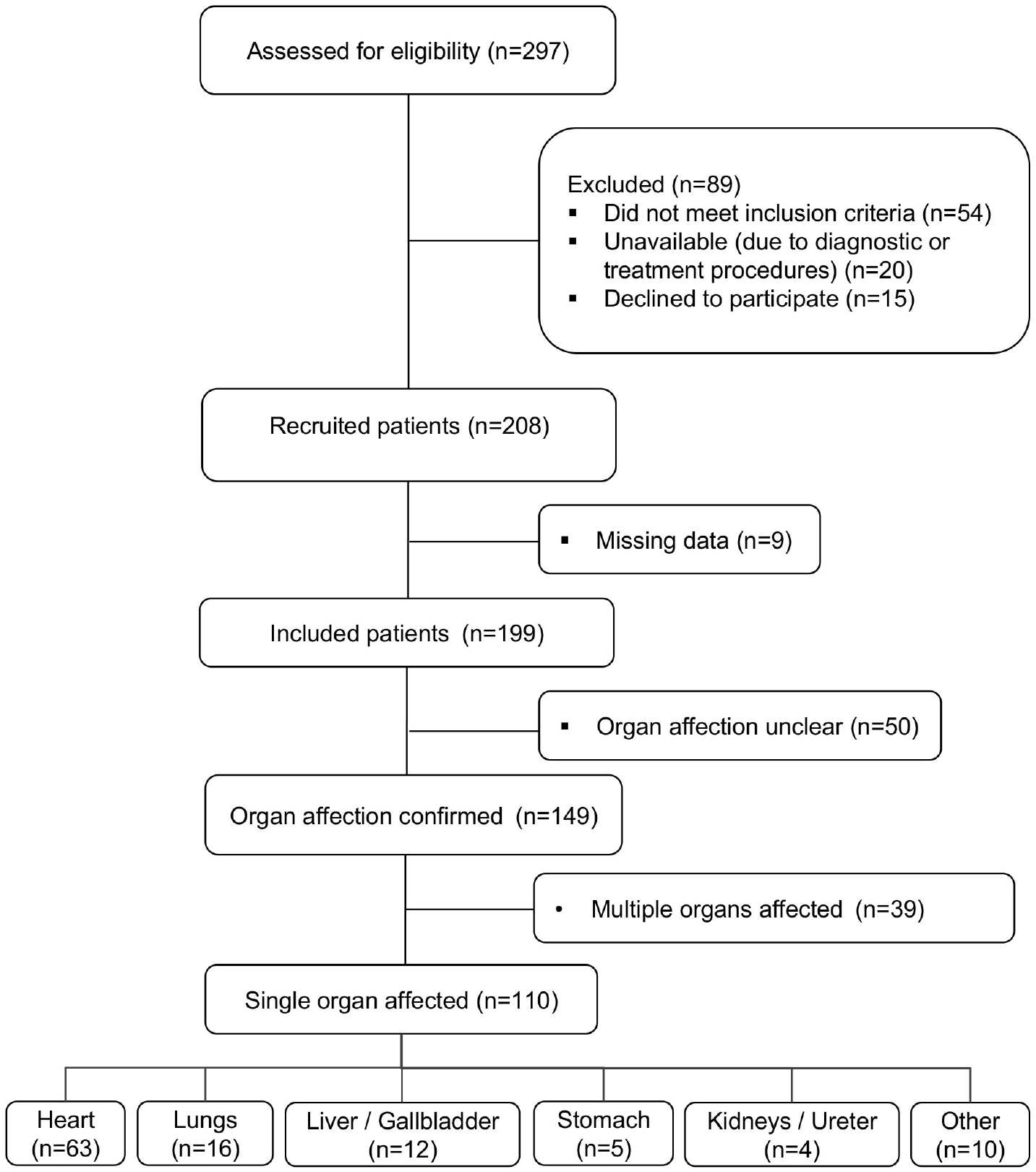
Flow-chart of the study.

We recruited 208 patients (86 women) for participation in our study. Nine drawings were lost due to technical failure of a tablet PC during the physical examination. The characteristics of the final study population can be found in **Table 2**.

**Table 2:** Demographics of the study population ^a,b^

|  |  |
| --- | --- |
| **Age (years), mean (SD)** | 57.26 (17.2) |
| **Age range, n (%)** |  |
| 18-39 | 34 (17.1) |
| 40-59 | 66 (33.2) |
| 60-79 | 80 (40.2) |
| 80+ | 19 (9.5) |
| **Women, n (%)** | 85 (42.7) |
| **The chief complaint, n (%)** |  |
| Chest pain | 88 (44.2) |
| Abdominal pain | 55 (27.6) |
| Dyspnea | 22 (11.1) |
| Other | 34 (17.1) |

<sup>a</sup>SD, standard deviation<sup>b</sup>Data are mean (SD) or n (%) unless otherwise specified.

### Procedures

All clinical data were collected by two of the authors (AA and NS), henceforth called examiners. AA is an internal medicine specialist, NS a physician with four years of training for internal medicine specialization. The examiners were fully informed about the study purpose and trained to do the physical examination for segmental signs and symptoms according to the protocol described below.

During recruitment, the examiners screened the emergency dashboard to identify patients who were referred to internal medicine specialists. They approached all eligible patients, informed them about the study, and obtained written informed consent.

The examination took place before primary pain treatment and before the final diagnosis was known to the examiner. It lasted between 7 and 15 minutes, depending on the patient’s compliance and interruptions by nurses and physicians. Directly after the physical examination, all findings were recorded on a tablet computer running the app “SymptomMapper” (see below).

### Categories of findings

The clinical findings we were interested in can be divided into three groups according to the ways they were recorded:

1. Distributed findings (i.e., those with a bodily pattern). These were spontaneous pain, allodynia, superficial hyperalgesia, deep hyperalgesia, superficial skin resistance, muscle resistance, defence, anisohydrosis, piloerection, vasomotor changes, herpes zoster, and resistance to passive movement of the limbs. Distributed findings were recorded by the examiners in the form of electronic drawings on a body template, thus capturing their exact location and extent.
2. Lateralized findings (i.e., those without a bodily pattern but with clear lateralization). These were anisocoria, glossy eye, eyelid separation, tense facial muscles, asymmetric posture, and reduced respiration movements. Lateralized findings were recorded by choosing from a list of all possible findings in conjunction with a side label (e.g., “glossy eye right”, “mydriasis left”, etc.).
3. Other findings. These were symptoms potentially related to organ-organ reflexes, namely nausea, vomiting, constipation, diarrhoea, meteorism, and urinary retention. Other findings were recorded by choosing from a simple list of all possible findings.

### Tablet computer and software application

All findings were recorded on Galaxy Note 10.1 (2014) tablet PCs with an electronic stylus based on inductive digitizing technology (Samsung, Seoul, South Korea). The tablets had a 10.1-inch touch screen with a resolution of 800×1280 pixels and were running Android 5.1.1 (Open handset alliance, Mountain View, CA, USA). The stylus was used for all data entry, hence allowing for a higher resolution while eliminating unwanted activation of the screen, for example, by the palm. Tablet and stylus were disinfected after every patient using wipe disinfection.

We used a modified version of the SymptomMapper app developed by our group to acquire electronic pain drawings [33]. Its usability for doctors and the reliability of its symptom drawing approach have recently been shown [33]. The app allowed the examiners to enter all findings from the bodily examination quickly. They could either draw distributed findings on a body template or choose from a list of lateralized or other findings. For the electronic drawings, examiners had a front and back view of the body available, and each newly added sign or symptom was displayed in a semi-transparent way and a different color.

### Bodily examination

Our approach to the bodily examination was based entirely on Hansen and Schliack [31] (p. 140-176). Its primary purpose was to check for the presence and record the extent and lateralization of pain and segmental signs and symptoms. We will start by describing how distributed segmental signs were collected, as this was done in the same way for different body regions (see below).

1. Visual inspection: The skin of the body region was visually inspected for the following signs:
  a. shingles (as a potential sign of Zoster reactivation);
  b. vasomotor changes, i.e. changes of skin color to the red, pale, or blue (as a sign of sympathetic reflexes);
  c. piloerection, i.e. any hair erection or “goosebumps” (as a sign of sympathetic reflexes).
2. Palpation: The body region was palpated with warm hands to test for the following signs of sympathetic reflexes, increased muscle tone or sensory disturbance:
  a. Anisohydrosis: The skin was observed and palpated for any local differences in the amount of sweating;
  b. Superficial hyperalgesia: The patient was informed that the examination could cause a little twinge. Then he or she was asked if the tip of a neurological examination needle (Healthstar, Lakewood, NJ, USA) passed vertically over the skin in long and slow strokes, caused a different sensation in any area;
  c. Deep hyperalgesia: For this test, folds of skin were held gently between thumb and index finger, or the region was tapped on. The test was considered positive if this procedure caused dull pain that lasted longer than in other parts of the body;
  d. Allodynia: The patient was asked if his or her clothes caused an unpleasant sensation somewhere on the body. Then he or she was asked if a medical cotton swab passed over the skin in long and slow strokes caused a different sensation in any area;
  e. Superficial skin resistance: This was tested by superficial palpation of the trunk skin using the palm with very soft pressure. If the examiner felt either resistance or a rubbery membrane in any area, the test was considered positive in this area;
  f. Muscle resistance: Deep palpation of the trunk wall was done on the front and back sides with the palm to detect the guarding of the trunk’s wall muscles.

The complete examination program was as follows. All steps were carried out in the exact order specified here.

1. Asymmetric posture: General inspection of the patients’ posture was carried out directly after entering the examination room to check side differences in muscle tone.
2. Pain and segmental symptoms: These were collected by asking patients the following questions:
  a. Do you have pain? Where exactly? Do you have a headache? In case the patient reported pain, the painful region was drawn.
  b. Do you have nausea? Did you vomit since the onset of symptoms?
  c. Do you have diarrhoea? Constipation?
  d. Do you feel that your abdomen is full of gases?
  e. Did you have any problem with urination since the onset of symptoms?
3. Head
  a. Special tests:
    i. Pupils: We tested for mydriasis, a sign of sympathetic activation, by equally exposing both eyes to light after instructing the patient to relax and look far away. The examiner used one hand to shadow the eyes and compared pupil diameter on both sides. This was repeated 3-5 times. In case of a striking side difference, the test was considered positive for mydriasis.
    ii. Eyes/eyelids: Eyelid separation and eye gloss, both signs of sympathetic activation, were assessed by comparing the visible area and gloss of both eyes. In case of a striking side difference, the more open and glossy eye was noted.
    iii. Tense facial muscles: This test checked for potential asymmetry of the facial features caused by side differences in muscle tone. It was considered positive when the upper lip was noticeably higher, the nasolabial fold deeper and the cheeks more retracted on one side than on the other. The test was repeated once under provocation by applying pressure on “Mussy’s point” between the two heads of the sternocleidomastoid muscle.
  b. Distributed segmental signs tested: zoster, vasomotor changes, piloerection, anisohydrosis, superficial hyperalgesia, allodynia, and superficial skin resistance.
4. Neck and chest The patient was examined on the front in a supine position and after freeing the chest. Then the back was examined with the patient sitting or lying on one side.
  a. Special tests The patient’s chest movement during inspiration and expiration was observed during the visual inspection, and any striking side differences noted as a sign of increased muscle tone.
  b. Distributed segmental signs tested: zoster, vasomotor changes, piloerection, anisohydrosis, superficial hyperalgesia, deep hyperalgesia, allodynia, superficial skin resistance, and muscle resistance.
5. Abdomen The patient was examined on the front in a supine position and after freeing the abdomen. Then the back was examined with the patient sitting or lying on one side.
  a. Special tests: Defence was examined by applying sudden deep palpation over the painful areas. If the examiner felt a reflex of the abdominal wall, it was considered positive.
  b. Distributed segmental signs tested: zoster, vasomotor changes, piloerection, anisohydrosis, superficial hyperalgesia, deep hyperalgesia, allodynia, superficial skin resistance, and muscle resistance.
6. Limbs
  a. Special tests: Passive movements of the joints were done to detect any resistance due to increased muscle tone.
  b. Distributed segmental signs tested: zoster, vasomotor changes, piloerection, superficial hyperalgesia, and allodynia.

### Patient selection

Medical reports of all recruited patients were followed up through Hannover Medical Schools’ electronic health records by three of the authors (NS, AA, and MM) to identify those patients with a definite diagnosis of visceral disease. All information regarding the acute complaint, previous diagnoses, and diagnostic procedures (electrocardiogram, laboratory, radiology, etc.) were reviewed, and the most likely aetiology for each patient discussed. Patients without a definite visceral diagnosis were excluded from further analysis (n=50). The remaining cases were divided into those where a single organ was affected (n=110) and those with multi-organ problems (n=39). Only the single-organ cases were included in the final analysis, and only organs with at least four patients were included in any organ-specific analyses.

### Data analysis

#### General considerations on lateralization

According to Hansen and Schliack, the majority of segmental signs are lateralized and appear on specific sides of the body defined by the innervation of the individual organs (see **Table 1**). In particular, the lateralization of signs for paired organs, such as lungs and kidneys, depends on which side is affected. Due to the nature of our study, it was not possible to conduct separate analysis for the left and right side in diseases of the lungs and kidneys/ureters. Furthermore, many of lung cases were bilateral affections. Information about the lateralization of segmental signs for lungs and kidneys/ureters is therefore of little value and only shown for the sake of completeness.

#### Lateralization and other findings

We extracted all lateralized findings (mydriasis, glossy eye, eyelid separation, tense facial muscles, asymmetric posture, and reduced respiration movements) and other findings (nausea, vomiting, constipation, diarrhoea, meteorism, and urinary retention) from SymptomMapper’s JavaScript Object Notation (JSON) files using a custom-written Python script (Python 2.7, Python Software Foundation, 2018). We then calculated for each organ and each finding the percentage of patients that had shown it. For lateralized findings, we calculated the percentage for each side individually treating front and back as one surface. Finally, we calculated the frequency of each finding averaged over all organs.

#### Distributed findings

Digital drawings from the app were converted to Nifti format (Neuroimaging Informatics Technology Initiative, 2017) with a custom-written Python script (Python 2.7, Python Software Foundation, 2018) and analyzed using tools from FMRIB Software Library (FSL) version 5.0 (FMRIB Analysis Group, Oxford University, UK). Figures were prepared using VINCI (“Volume Imaging in Neurological Research, Co-Registration and ROIs Included”) 4.86.0 (Max Planck Institute for Metabolism Research, Cologne, Germany) and GNU Image Manipulation Program version 2.8.16 (GIMP, The GIMP Team). In all analyses, the front and back were treated as one surface.

First, to derive the bodily distribution of all segmental signs, all distributed signs were superimposed and the result binarized. In the resulting map, a pixel of value one on the body template meant that at least one sign had been found at that particular point on the body in that particular patient. Binarization meant that we disregarded the number of signs that each patient showed and instead only considered their bodily location. The organ-specific maps were color-coded using a heat map color palette, where warmer colors indicated a higher degree of overlap between patients.

We then analyzed distributed signs individually to assess the segmental distribution for each sign according to the segmental scheme of Hansen and Schliack, which is largely based on Head’s [12,31]. To do so, we calculated for each segment the percentage of the segment covered by the sign. This was done by dividing the pixel count by the total number of pixels of the respective segment. Only segments with at least five per cent coverage were included. To assess the lateralization of findings, we further divided segments into left and right body halves, calculating the percentage for each of them. This resulted in a list of half segments covered by each sign. Finally, we calculated for each organ the average number of segmental signs per half segment and the frequency of each sign averaged over all organs.

Spontaneous pain was analyzed in the same way, but separately from all other signs.

## Results

### The overall frequency of signs and symptoms

Of the 110 patients in our final sample, 85 had spontaneous pain, while 81 showed at least one segmental sign and 52 at least one segmental symptom. On average, each patient had 1.80±1.86 (mean±SD) segmental signs and 0.77±1.00 segmental symptoms. The most frequent signs and symptoms are shown in **Table 3**.

**Table 3:** Frequency of segmental signs and symptoms in our patient sample (n=110).

| **Segmental signs** | **n** |
| --- | --- |
| Superficial hyperalgesia<br>(Head's zone) | 46 |
| Muscle resistance | 39 |
| Mydriasis | 37 |
| Defence | 13 |
| Deep hyperalgesia<br>(Mackenzie's zone) | 13 |
| Superficial skin resistance | 12 |
| Tense facial muscles | 11 |
| Vasomotor changes | 10 |
| Glossy eye/wide eyelid | 8 |
| Asymmetric posture | 7 |
| Reduced respiration movements | 3 |
| Allodynia | 2 |
| Piloerection | 1 |
| Anisohydrosis | 1 |
| Zoster | 0 |
| *At least one segmental sign* | 84 |
| **Segmental symptoms** | **n** |
| Nausea | 45 |
| Vomiting | 18 |
| Diarrhoea | 10 |
| Meteorism | 8 |
| Constipation | 5 |
| Urinary retention | 0 |
| *At least one segmental symptom* | 52 |
| **Spontaneous pain** | **85** |

### Frequency of lateralization signs and symptoms

All lateralization signs and segmental symptoms are shown in **Suppl. Fig. 1**. As predicted, the majority of lateralization signs were detected ipsilateral to the affected organ for the unpaired organs heart, stomach and liver/gallbladder. The most striking finding was the high number of patients showing ipsilateral mydriasis as a sign of unilateral sympathetic activation. This lateralization was 100 per cent ipsilateral for diseases of the liver/gallbladder (5 right vs 0 left), 100 per cent ipsilateral for stomach diseases (1 left vs 0 right), and 83 per cent ipsilateral for heart diseases (15 left vs 3 right).

### Segmental signs and spontaneous pain in individual patients

Bodily maps of segmental signs and spontaneous pain for a selection of individual patients are shown in **Figure 2**. These cases reflect the entire bandwidth of segmental signs encountered in patients presenting to the emergency room. Their primary diagnoses and demographic information is summarized in **Suppl. Tab. 1**.

**Figure 2:**
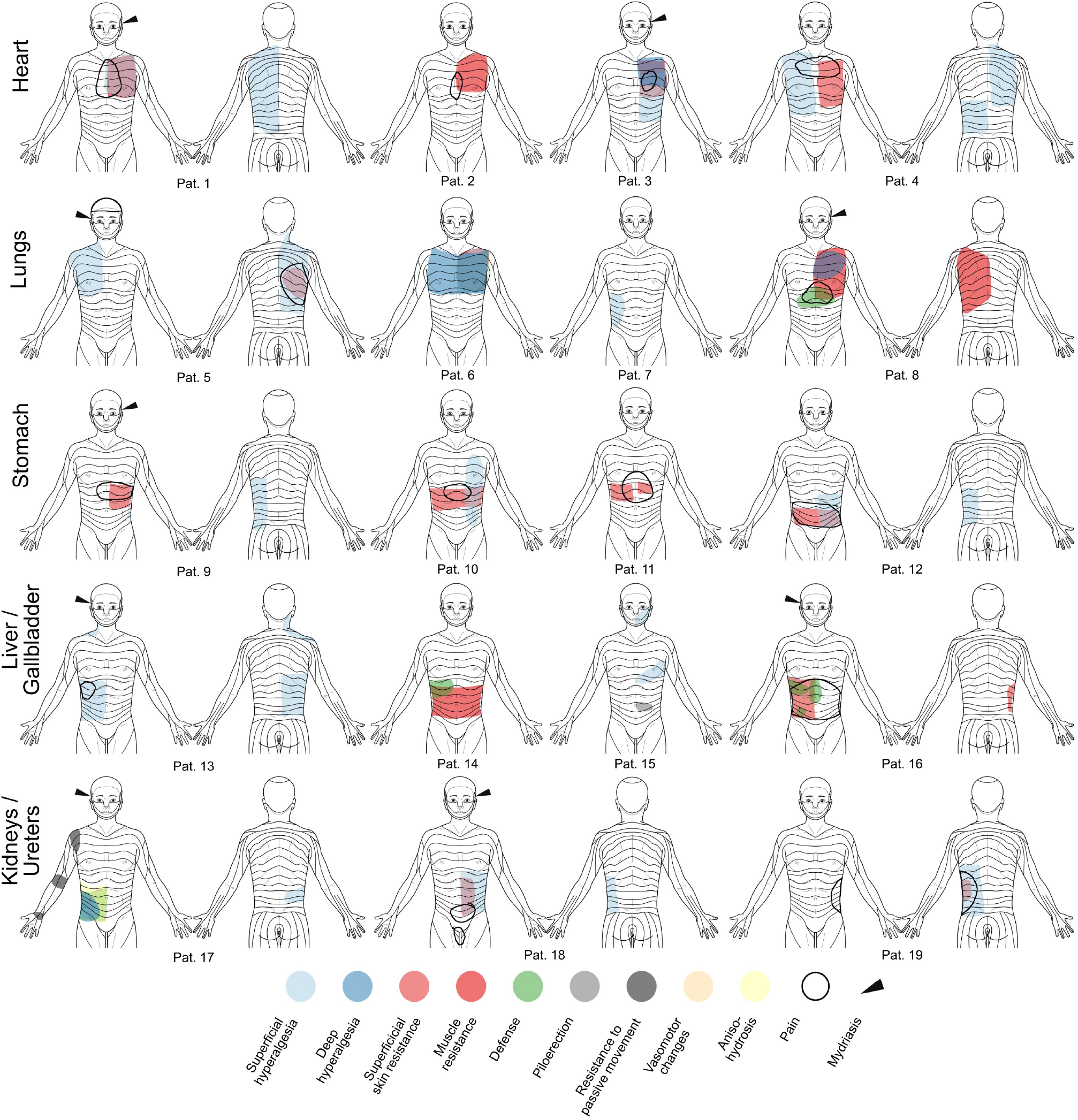
Segmental signs and spontaneous pain in individual patients with acute visceral diseases. For primary diagnoses and demographic information see **Suppl. Tab. 1**. The cases shown here reflect the entire bandwidth of segmental signs encountered in patients presenting to the emergency room. It ranges from “textbook cases” (e.g. patients 2, 3, 6, 9, 13, 18, and 19), where segmental signs alone allow for a preliminary diagnosis, to those, where segmental signs are hardly helpful or even misleading (e.g. patients 7, 12, and 15).

### Bodily maps and segmental patterns of distributed signs

Average bodily maps of all distributed segmental signs are shown in **Figure 3**, while **Figure 4** contains detailed segmental information concerning the distribution of the individual signs and spontaneous pain. In general, the observed distributions of segmental signs were largely consistent with those reported by Hansen and Schliack [31]. The only exception was the lungs, which showed a more widespread distribution than predicted. Concerning lateralization, segmental signs from the unpaired organs showed a clear side difference with more signs appearing ipsilateral to the affected organ, thus giving support to the “side rule” (cf. **Table 1**).

**Figure 3:**
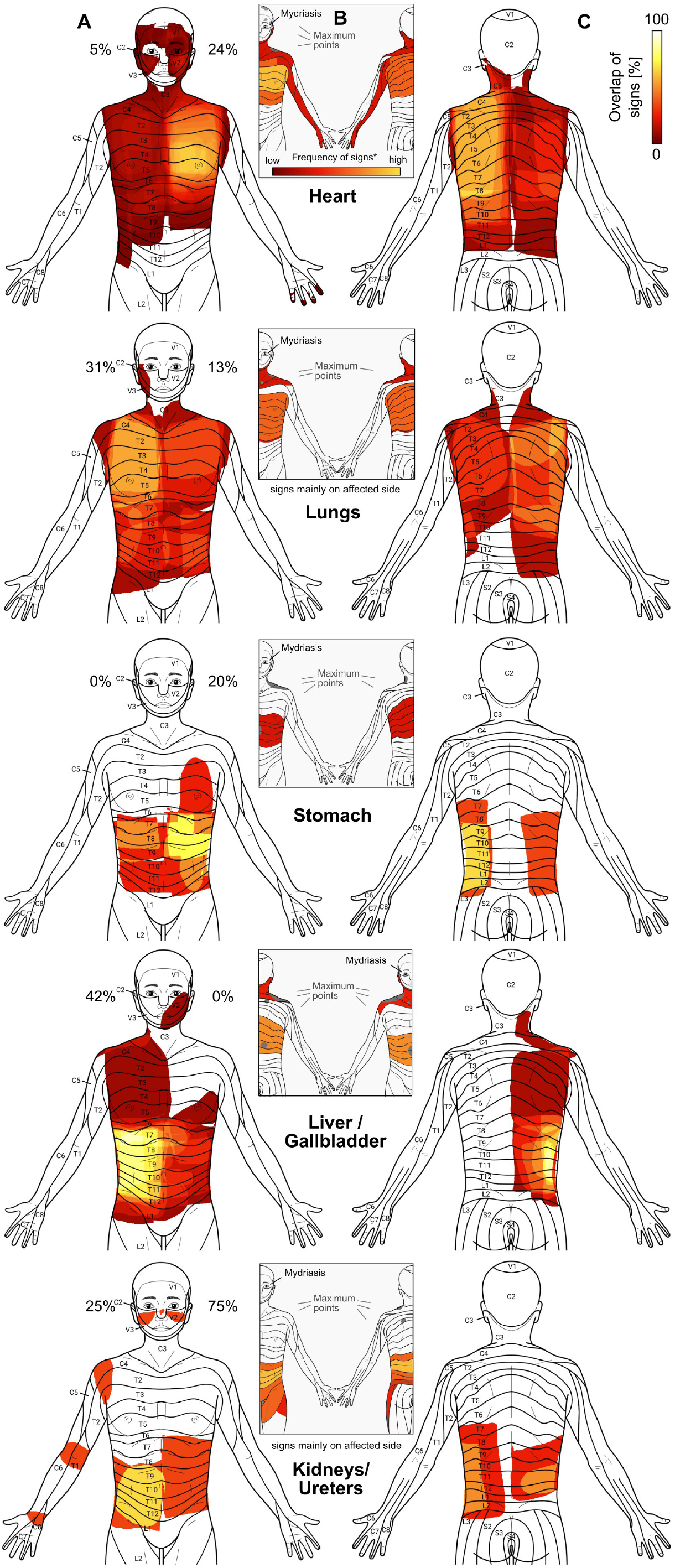
Distributed segmental signs in acute visceral diseases. Warmer colours reflect higher overlap of signs between patients. Columns A and C show a joint map (front and back) of all distributed segmental signs. The inserts in column B show the segmental distributions for each organ as reported by Hansen and Schliack [31] for comparison. Percentage values at the sides of the head indicate the frequency of unilateral mydriasis in affections of the respective organ. In general, the bodily patterns of segmental signs were largely consistent with those reported by Hansen and Schliack except for the lungs, where the signs were much more widespread. The distribution of the signs from the unpaired organs showed a clear side difference with more signs appearing ipsilateral to the affected organ as predicted by the “side rule”. For the lungs and kidneys/ureters, however, this rule could not be tested, since results for these organs reflected a mixture of left, right and bilateral affections.

**Figure 4:**
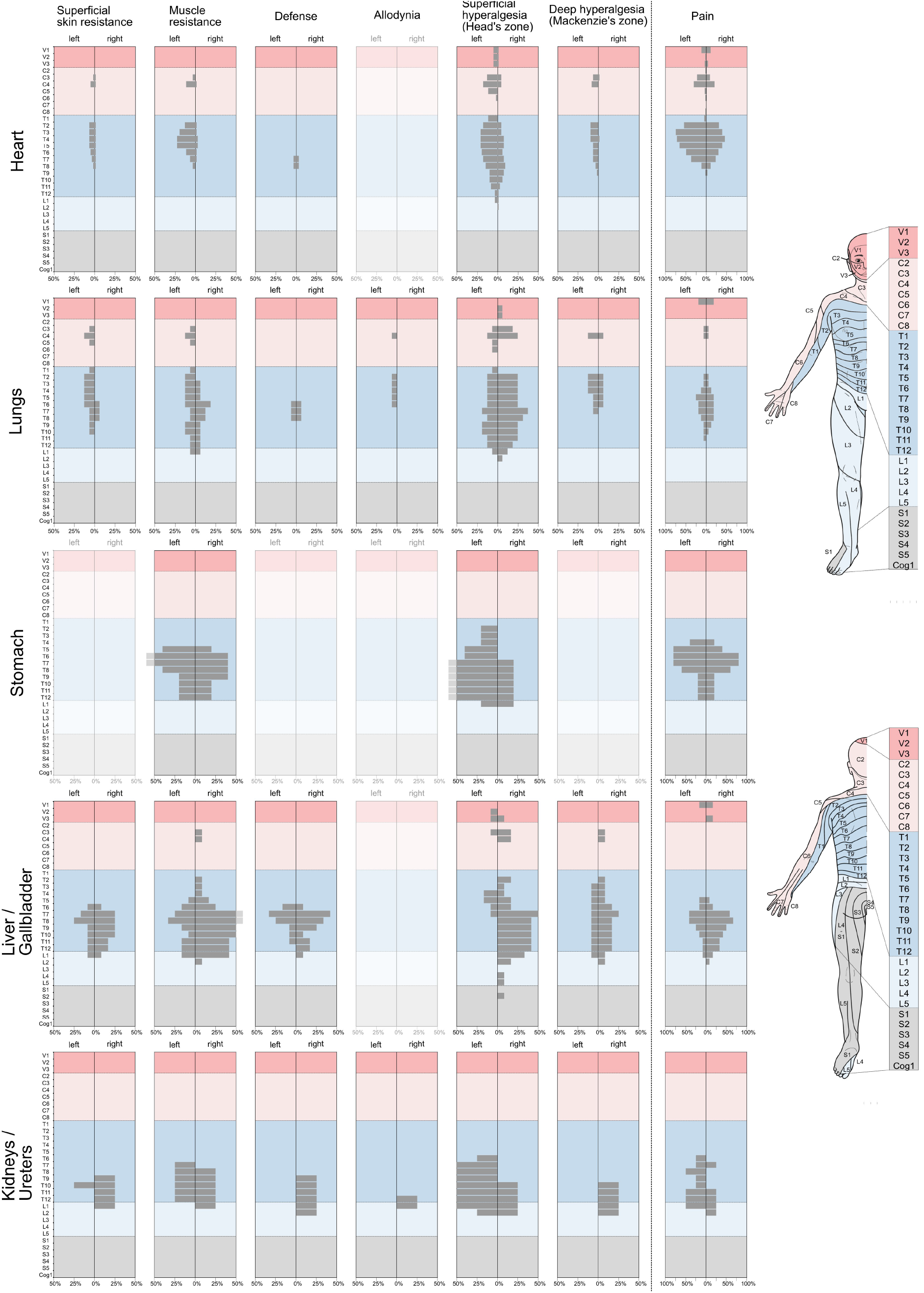
Segmental patterns and lateralization of segmental signs and pain in acute visceral diseases. Each row contains the results for an individual organ, while columns represent the different most common segmental signs and pain respectively. Please note the different axis scaling for the latter. For the sake of clarity, segmental sections have been colour-coded according to the body template shown on the right. Within-organ comparison shows that the different segmental signs but also spontaneous pain have a similar segmental distribution. Between organs, these distributions show considerable overlap. Superficial hyperalgesia (Head’s zone) exhibits the greatest spread in terms of segments. Regarding lateralization, segmental signs for the unpaired organs mostly obeyed the side rule, according to which signs should appear on the body half, where the organ is located. Pain differed markedly in that respect and instead showed a rather symmetric pattern. For the paired organs lungs and kidneys/ureters, the rule could not be tested since results for these organs reflected a mixture of left, right and bilateral affections.

For diseases of the heart, segmental signs were mostly located in the thoracic segments and to a lesser extent in cervical segments. Superficial hyperalgesia (Head’s zone) was also detected in the trigeminal segments. The maximum of the averaged signs was in the T3-T5 region as predicted by Hansen and Schliack (**Figure 3B**). In terms of lateralization, all distributed signs were strongly left-dominated with deep hyperalgesia (Mackenzie’s zone) showing complete left-lateralization (**Figure 4**). Defence and allodynia were rare and non-existent, respectively, in heart patients.

For diseases of the lungs, segmental signs were very widespread and covered a range from V2 to L2. Signs were generally less focused than for the heart, and no clear maximum was discernible. In this, the distribution deviated from Hansen and Schliack’s, who give T9 as the lower margin of segmental signs in lung diseases. As was to be expected due to the mixture of left, right and bilateral organ diseases, no lateralization could be seen.

For the stomach, the segmental distribution was almost strictly thoracic from T2 to T12 with a maximum at T6-9 on the front and on the back. Similar to the heart, superficial hyperalgesia for the stomach was lateralized to the left as predicted by the side rule. The comparison with Hansen and Schliack showed that the maximum of signs in T6-9 fell into the expected range in the front view. On the back, however, there was only a partial overlap with Hansen and Schliack predicting higher thoracic segments than were found in our study.

The similarity with Hansen and Schliack’ results was much higher for patients with liver/gallbladder diseases. Here, segmental signs showed a largely thoracic distribution but with the characteristic shoulder presentation in segments C3-C5. In terms of lateralization, muscle resistance, defence, superficial and deep hyperalgesia were predominantly right-lateralized with superficial hyperalgesia (Head’s zone) showing almost complete right-lateralization.

Finally, segmental signs of the kidneys had the narrowest distribution starting at T6 and extending down to L2, once again showing a rather high similarity with the predicted distribution by Hansen and Schliack.

### Comparison of spontaneous pain and segmental signs

The segmental distributions of spontaneous pain and segmental signs are shown in **Figure 5**. It is evident that spontaneous pain differed markedly from segmental signs. It spanned fewer segments but extended to the head region (V1) in heart, lung, and liver/gallbladder diseases. Furthermore, spontaneous pain was much less lateralized than segmental signs but rather localized in the body midline.

**Figure 5:**
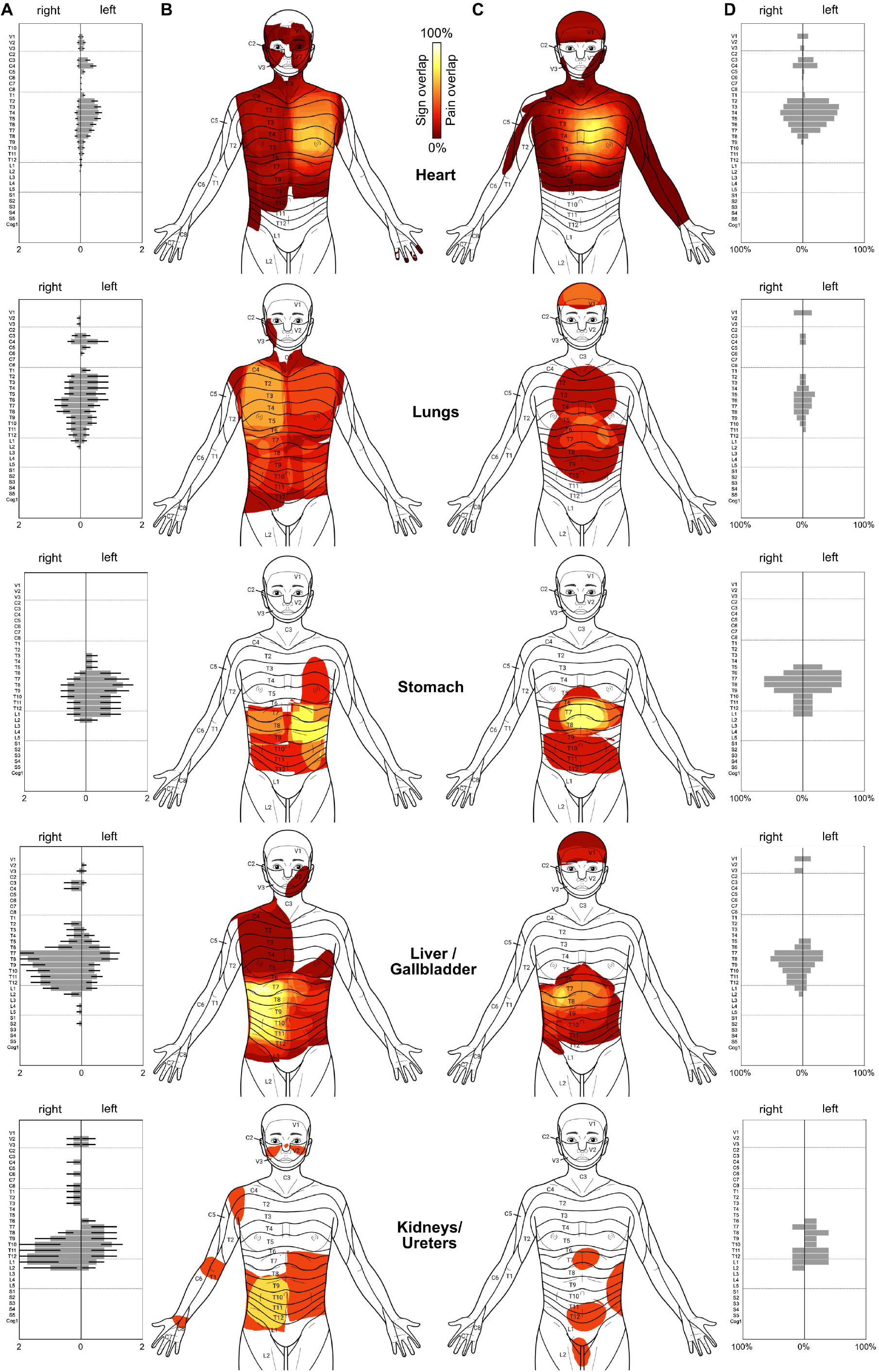
Direct comparison between segmental signs (A,B) and spontaneous pain (C,D) in acute visceral diseases. Column A shows the average number of segmental signs per segment for the individual organs, column B the joint distribution of segmental signs (cf. Figure 2). The symptom of spontaneous pain is shown as average pain distributions in column C and their exact segmental content in column D. In general, the pain was less widespread and showed much weaker lateralization than segmental signs in the unpaired organs (heart, stomach, liver/gallbladder).

## Discussion

In this study, we investigated bodily patterns and lateralization of segmental signs and spontaneous pain in acute visceral diseases. We derived average distributions of spontaneous pain and segmental signs for the heart, lungs, stomach, liver/gallbladder, and kidneys/ureters by combining digital pain drawing technology and a structured 10-minute bodily examination in patients presenting to the emergency room. We extracted precise information on the segmental content and lateralization and compared the results to the slightly outdated but authoritative German work of Hansen and Schliack [31]. Although purely descriptive by design, our study is the first in the English language to give a detailed account of simultaneously collected segmental signs and symptoms for visceral diseases in the clinical setting.

### Lateralization of segmental signs

The importance of the lateralization of segmental signs lies in the possibility to quickly identify the affected body side, i.e. the side hosting the affected organ (**Table 1**). Such side rule may be useful in the differential diagnosis, such as in differentiating gastritis from hepatitis, or pancreatitis, or acute coronary syndrome from a pulmonary embolism, or esophagitis. Due to our study design and the very mixed patient sample, our results regarding lateralization are limited to the heart, stomach, and liver/gallbladder. Although segmental signs of the lungs and kidneys/ureters are also expected to be found ipsilateral to the affected side, a separate analysis for the individual sides was not possible for these organs due to the limited number of cases, many of whom showed bilateral affections.

Although our data were not analyzed prospectively, it seems safe to say that for heart, stomach, and liver/gallbladder they support the findings of Hansen and Schliack that segmental signs appear ipsilateral to the affected organ [31]. While this was evident for the averaged bodily maps of distributed signs, we also found an ipsilateral occurrence of mydriasis, a finding rarely raised outside the neurological setting. It results from a reflex mediated by the ciliospinal centre, which conducts impulses from the entire body to the sympathetically innervated dilator pupillae muscle [26] (p.271) and has first been described to occur in affections of the lungs [34] and the heart [35] more than a hundred years ago.

We found right-sided mydriasis in 42 per cent of our liver/gallbladder patients and not a single case of left-sided mydriasis. For the heart, mydriasis was less frequent (24 per cent left-sided vs five per cent right-sided), yet this means that patients showing the sign, had it on the ipsilateral side in almost 83 per cent of the cases. Hansen and Schliack report qualitatively similar but generally higher numbers for mydriasis. In their sample of 28 heart patients, 27 (i.e. 96 per cent) had mydriasis that was ipsilateral in 26 patients (i.e. 96 per cent). In 56 liver/gallbladder patients, 54 (i.e. 96 per cent) had mydriasis, of which 50 cases (i.e. 93 per cent) were ipsilateral (i.e. right-sided).

The generally higher numbers of mydriasis in heart diseases found in Hansen and Schliack’s work may be explained by the fact that these authors used dark adaptation and infrared photographs in many of their patients, while our examiners were restricted to visual inspection under normal light. Clinicians interested in this phenomenon should consider using a portable infrared pupilometer.

### Localization and distribution of segmental signs

A subset of the findings collected in our study was further analyzed to extract detailed segmental information. We called this group of findings “distributed signs”. It comprised a number of somatosensory (superficial and deep hyperalgesia, allodynia), somatomotor (superficial skin resistance, muscle resistance, defence), and visceromotor signs (vasomotor changes, piloerection, anisohydrosis). Of these, superficial hyperalgesia (i.e. Head’s zones), muscle resistance, defence, and deep hyperalgesia (i.e. Mackenzie’s zones) were the most frequently observed in our sample of patients, while others, like allodynia, piloerection, anisohydrosis, or zoster were exceedingly rare.

Once again, we found our findings to support those by Hansen and Schliack in that there was a close similarity between the original maps of segmental signs by these authors and our averaged maps of all distributed signs (**Figure 3**). For a prospective evaluation, however, future studies should aim to quantify this similarity, e.g. by using spatial similarity measures.

Several groups have studied individual segmental signs or groups of signs since the days of Hansen and Schliack. For example, Nicholas and colleagues found that patients with myocardial infarction showed characteristic paravertebral soft tissue changes readily detected by palpation. Compared to a control group, patients with myocardial infarction had a significantly higher incidence of increased firmness, warmth, ropiness, oedematous changes, and heavy musculature, almost entirely confined to segments T1-4 [36]. In a follow-up three years after the infarction, these signs had regressed in the majority of patients [37]. Vecchiet and colleagues investigated superficial and deep hyperalgesia of the L1 segment in patients after renal/ureteral calculosis finding that pain thresholds were lower on the affected side with respect to both the contralateral side and control thresholds recorded in healthy subjects [38].

For the gallbladder, Stawowy and colleagues found that all patients with acute cholecystitis reported referred pain in the epigastrium and under the right curvature. Segmental signs inside this area were quantitatively evaluated using von Frey hairs, warm and cold metal rollers, and a constant current stimulator to test for the different forms of hypersensitivity or allodynia. The authors reported that 20 per cent of the patients showed hypersensitivity or allodynia to mechanical, 53 per cent to cold, 40 per cent to warmth, and 63 per cent to electrical stimulation [39]. The same authors reported, that 50 to 56 per cent of patients with acute appendicitis showed segmental signs over the right abdominal quadrant, with the maximum located approximately at McBurney’s point [40]. These findings were recently confirmed by Roumen and colleagues, who reported that 39 per cent of patients with acute appendicitis demonstrated at least one segmental sign (hyperalgesia, hypoesthesia, altered cool perception, positive pinch test) over the lower right abdomen [27]. Finally, a large number of smaller studies and case reports have been published, many of them from the osteopathic profession, and have been reviewed by Beal [41].

### Segmental signs vs spontaneous pain

The majority of our patients with visceral diseases reported spontaneous pain (**Table 3**). With 85 per cent of the cases, it was by far the most frequent finding, followed by superficial hyperalgesia (46 per cent), nausea (45 per cent), and muscle resistance (39 per cent).

Many textbooks assign pain location a discriminative role in the differential diagnosis. However, such predictive power of pain location has been a matter of debate for decades [9-11]. Here, we found by direct comparison of spontaneous pain and segmental signs that the two were rather dissimilar in their bodily pattern and segmental distribution (**Figure 5**). Irrespective of the affected organ, spontaneous pain was less widespread than segmental signs, i.e. it included fewer segments. Furthermore, spontaneous pain appeared mostly in the body midline, thus lacking the diagnostically relevant ipsilateral distribution seen in the majority of segmental signs. As **Figure 5** shows, patients with lung, stomach, and liver/gallbladder diseases all showed spontaneous pain in the epigastric region (T5-9), thus rendering this symptom unsuitable for differential diagnosis.

The substantial differences found between pain and segmental signs regarding their location and lateralization underline the importance of making a clear distinction between visceral pain, referred pain, (referred) hyperalgesia, and other segmental signs. While primary visceral pain is a poorly defined, midline sensation, it starts to be referred or ‘transferred’ to somatic structures, when it persists for several minutes or longer [25,42]. These somatic structures can include skin, subcutaneous tissue, and muscle, and are characterized by an overlapping segmental innervation with that of the diseased organ. Pain referral to the skin or muscle is what triggers the additional somatosensory, somatomotor, or autonomic sign seen in our study.

Despite the purely descriptive design of this study, our results regarding the benefit of using spontaneous pain or segmental signs seem to favor the latter over the former. Future studies should test this in a prospective way.

### Limitations

Our study has several limitations that need to be discussed. Firstly, our patient sample was relatively small, as we could only analyze about half of the patients we had included. There were two reasons for this. On the one hand, about one-quarter of our patients had to be excluded from the analysis, since it left the hospital without a confirmed final diagnosis. This is due to the unique situation in the emergency room, where the vital role of the specialist is to rule out life-threatening conditions. A further quarter of the remaining patients had to be excluded from the analysis because it suffered from diseases affecting multiple organs. Secondly, while we took great care to include only patients with single-organ problems, it is likely that affections of other organs were present but overlooked in some of our patients. Thirdly, findings collected by means of palpation are naturally more subjective than, e.g. laboratory results. While there are ways to measure segmental signs more quantitatively, we did not do so to keep the examination time to an absolute minimum as required by the clinical setting.

### Conclusions

The present study underlines the usefulness of including segmental signs in the bodily examination of patients with acute medical problems. Segmental information and lateralization may help to narrow the number of possible causes before more sophisticated test results can be obtained. Clinicians interested in segmental diagnosis may start by testing for the three most frequent signs, i.e. superficial hyperalgesia (Head’s zones), muscle resistance, and mydriasis. They need nothing more than their hands and a neurological examination needle.

## Data Availability

All data are available from the authors upon reasonable request.

## Acknowledgments

The authors would also like to thank Christoph Duesberg, former Head of the Central Emergency Room at Hannover Medical School, for his support in planning and carrying out the study.

